# Spinal Cord Infarction Outcomes with a Focus on Thrombolysis: A Nationwide Database Study

**DOI:** 10.1101/2024.07.08.24310117

**Authors:** Ali Al-Salahat, Danielle B. Dilsaver, Ali Bin Abdul Jabbar, Saif Bawaneh, Abhishek Singh

## Abstract

**Background:** Spinal cord infarction (SCI) is a rare and challenging condition to diagnose. The literature lacks large studies examining the characteristics and outcomes of SCI. Treatments, such as thrombolysis, which are considered standard of care in cerebral stroke, have not been previously studied in SCI. The primary aim was to determine outcomes of hospitalizations with SCI in the United States (US) by assessing inpatient mortality and all-cause 30-day and 90-day readmissions. After adjusting for age and comorbidity burden, we analyzed if these outcomes differed based on thrombolysis, and the most common concomitant conditions, such as aneurysms, dissections and sepsis.

**Methods:** Hospitalizations of patients over 18 years of age with SCI were abstracted from the 2016-2021 Nationwide Readmissions Database (NRD). We estimated logistic and lognormal regression models to evaluate whether outcomes differed based on thrombolysis, or concomitant aneurysm, dissection, or sepsis. Multivariable models adjusted for age and comorbidity burden.

**Results:** An estimated 12,548 hospitalizations involved patients diagnosed with SCI; of those, 21.43% were specifically for SCI. Other common reasons for SCI hospitalization included aneurysms or dissections (22.22%) and sepsis (9.11%). The average LOS was 13.09 days (95% CI: 12.61-13.59), with an inpatient mortality rate of 14.66% (95% CI: 13.74-15.59). Age and comorbidity burden significantly impacted inpatient mortality, LOS, and hospital costs. Hospitalizations of patients with SCI were predominately at large bed size facilities (72.36%, Weighted N: 9,079) and metropolitan teaching hospitals (86.57%, Weighted N: 10,862). Thrombolysis was associated with lower odds of 90-day readmissions (adjusted OR=0.35, p-value= 0.009) and no significant difference in mortality. Concomitant sepsis significantly increased mortality, LOS, cost, and readmission odds.

**Conclusions:** This study highlights factors that impact the outcomes of SCI on a large scale. Additionally, the findings uncover the association between thrombolysis and better long-term outcomes in SCI. This paves the way for further investigation into the role of thrombolysis in SCI.

## Introduction

Spinal cord infarction (SCI) accounts only for 0.3 to 1% of all strokes, and 6% of all acute myelopathies^1,2^. The most frequently affected spinal cord segments are, in descending order, thoracic, cervical and the conus medullaris^2,3^. Clinical presentation of SCI consists of an acute myelopathy, accompanied by sphincteric and autonomic dysfunction, and back pain^2,3^. Previous work has reported a median age of SCI onset between 52 and 60 years^3^. The diagnosis of SCI can be challenging to confirm, and misdiagnosis is common^2^. Zalewski et al. described diagnostic criteria that includes clinical, radiological and CSF findings to aid in the diagnosis of SCI in clinical practice and for research purposes^3^. Etiologically, SCI can be classified into spontaneous and peri-procedural. Spontaneous SCI is often attributed to atherosclerosis and vascular risk factors, whereas aortic repair surgeries contribute to the majority of peri-procedural SCIs^3–5^. The most reported risk factors for SCI include hypertension, peripheral vascular disease and advanced age^3,4^. Additionally, spontaneous SCI has been reported in association with degenerative spine disease, owing to fibrocartilaginous embolization^9,10^. Regarding the risk of peri-procedural SCI with open versus endovascular aortic repairs, studies showed conflicting results^6,7^. A meta-analysis by Gaudino et al. showed a rate of SCI of 5.7% for open repair and 3.9% for endovascular repair^8^.

The management of SCI is limited, lacking large randomized controlled trials. The role of thrombolysis in SCI is not established, with only a few case reports and series that showed potential beneficial effects^12,13^. Lumbar cerebrospinal fluid drains have been proven to aid in the prevention of SCI in association with aortic repair surgeries^14^. Studies have shown that recovery after SCI can be slow and gradual over months to years^2,11^. Furthermore, the functional recovery is much worse after peri-procedural SCI compared to spontaneous SCI^2^. This is attributed to the extent of spinal cord segments involved in peri-procedural SCI^2^. The 1-year and 5-year mortality rates of SCI were shown to be around 7% and 20.9%, respectively^15^. In addition, no significant difference was found in the survival of patients affected by SCI due to different etiologies^15^. The literature lacks large studies analyzing characteristics and outcomes of SCI. Given the rarity of SCI, standard treatments for cerebral stroke, such as thrombolysis, have not been previously studied. In this study, we aim to assess the characteristics and outcomes of SCI in a large sample representative of the United States, using the Nationwide Readmissions Database (NRD). The advantage of using the NRD as the data source for our study is that readmission rates are considered good indicators for long-term morbidity in cerebrovascular disease and stroke. Additionally, it allows us to capture as many hospitalizations as possible when it pertains to rare conditions and exposures, such as SCI and thrombolysis.

## Methods

### Data Source and Identification

Hospitalization data were abstracted from 2016 through 2021 NRD. The NRD is an all-payer, administrative inpatient care database sponsored by the Agency for Healthcare Research and Quality (AHRQ) and estimates approximately 35 million yearly discharges in United States^16^. We identified hospitalization of patients with a diagnosis of SCI using *International*

*Classification of Diseases, Tenth Revision Codes* (ICD-10: G95.11). Hospitalizations were excluded if the patient was younger than 18 years. Creighton University acknowledged this study as Not Human Subjects Research (InfoEd Record Number: 2004608); the NRD is HIPAA-compliant and publicly available. This study followed the STROBE reporting guidelines^17^.

### Study Outcomes

The primary aim was to assess the characteristics of hospitalizations of patients with SCI in a nationally representative sample. First, we described reasons for SCI hospitalization and the descriptive characteristics of the SCI hospitalization cohort. Then, we assessed whether inpatient mortality and length of stay (LOS) were associated with age and comorbidity burden. Lastly, we evaluated whether hospitalization outcomes varied 1) by those who received thrombolysis, 2) those who experienced an aneurysm or dissection, and 3) those that carried a sepsis diagnosis. Hospitalization outcomes included inpatient mortality, LOS, hospital cost, discharge disposition (routine, transfer to short-term hospital, transfer to other facility), 30-day all-cause readmission, and 90-day all-cause readmission.

Most frequent reasons for hospitalization were identified using the primary ICD-10 diagnosis code. ICD-10 diagnosis and procedures codes were used to identify SCI hospitalizations with receipt of thrombolysis (ICD-10-CM; Z92.82, ICD-10-PCS: 3E03017, 3E03317). ICD-10 diagnosis codes were used to identify SCI hospitalizations of patients with an aneurysm/dissection (ICD-10-CM: I71, I72) or sepsis (ICD-10-CM: A40, A41). Hospital cost was inflation adjusted to July-2021 US dollars^18^. For readmission analysis, if a patient died, the hospitalization was excluded to allow for complete post-discharge follow-up. Additionally, index hospitalizations where the patient was discharged in December were excluded for 30-day readmission analysis and index hospitalizations where the patient was discharge in October, November, or December were excluded for 90-day readmission analysis to allow complete post-discharge follow-up since the NRD does not track patients year-to-year.

### Descriptives

We extracted descriptives for each hospitalization; descriptives were age, biological sex (male, female), primary payer (Medicaid, Medicare, private, other, unknown), income quartile (I, II, III, IV, unknown), whether the hospital admission was elective, whether the hospital admission occurred on a weekend, facility bed size (small, medium large), facility location-teaching status (metropolitan non-teaching, metropolitan teaching, non-metropolitan), comorbidity burden, and National Institutes of Health Stroke Scale (NIHSS) score. Comorbidity burden was quantified by the Charlson Comorbidity Index (CCI); a score developed for healthcare research to assess patient risk^19^. We identified the following Charlson comorbidities: myocardial infarction, congestive heart failure, peripheral artery disease, stroke, dementia, chronic obstructive pulmonary disease, rheumatic disease, peptic ulcer, liver disease (mild, severe), diabetes (uncomplicated, complicated), renal disease (mild, severe), malignancy, metastatic tumor, human immunodeficiency virus, and acquired immunodeficiency syndrome. We also identified whether the patient underwent an aortic repair during the hospitalization using ICD-10 procedures codes (Open Approach: 02QX0ZZ, 02QW0ZZ, 04Q00ZZ; Percutaneous Approach: 02QX3ZZ, 02QW3ZZ, 04Q03ZZ).

### Statistical Analysis

Continuous descriptive were presented as median and interquartile range and categorical descriptives were presented as weighted frequency and percent. Logistic regression models were estimated to evaluate whether inpatient mortality varied by age and CCI. Log-normal models were estimated to evaluate whether LOS varied by age and CCI.

Next, to assess whether inpatient mortality, discharge disposition, and 30-day and 90-day all-cause readmissions differed by 1) the receipt of thrombolysis, 2) whether the patient experienced a dissection or aneurysm, and 3) whether the patient suffered from sepsis, we estimated respective logistic regression models. Likewise, we estimated log-normal models to evaluate whether LOS and hospital cost differed by 1) thrombolysis, 2) dissection or aneurysm, and 3) sepsis. Multivariable regression models were estimated that controlled for age and comorbidity burden.

The functional form of continuous covariates was evaluated via restricted cubic splines with knots at the 10^th^, 50^th^, and 90^th^ percentiles^20^. The NRD sampling design was accounted for in all analysis; two-tailed p < 0.05 indicated statistical significances.

## Results

In the United States from 2016 through 2021, there were an estimated 12,548 (95% CI: 12,085-13,010) hospitalizations where the patient carried a SCI diagnosis. Of those, 21.43% (Weighted N: 2,689, 95% CI: 2,526-2,853) were hospitalizations specifically for SCI. Other frequent reasons for hospitalization included hospitalizations for an aneurysm or dissection (22.22%, Weighted N: 2,537, 95% CI: 2,320-2,754) and sepsis (9.11%, Weighted N: 1,143, 95% CI: 1,131-1,327). **Table 1** presents hospitalization descriptives. Notably, there were no hospitalizations of patients with SCI where the patient underwent a percutaneous aortic repair. Hospitalizations of patients with SCI were predominately at large bed size facilities (72.36%, Weighted N: 9,079) and metropolitan teaching hospitals (86.57%, Weighted N: 10,862). Patients were primarily insured by Medicare (52.43%, Weighted N= 6,578). Common comorbidities were peripheral artery disease (37.58%, Weighted N=4,715), chronic obstructive pulmonary disease (23.00%, Weighted N: 2,886), congestive heart failure (22.22%, Weighted N = 2,788), and acquired immunodeficiency syndrome (21.46%, Weighted N = 2,693).

**Table 1.**
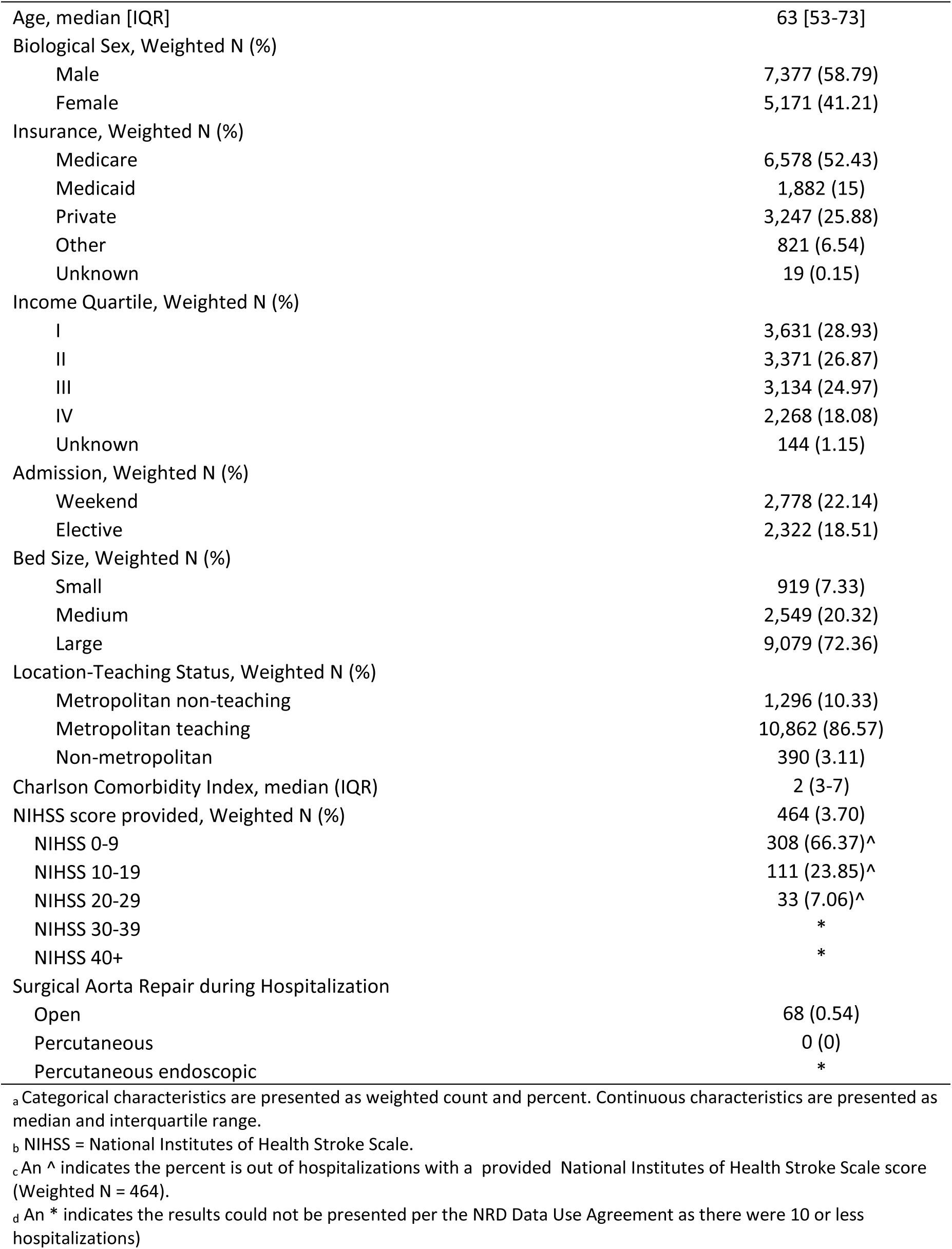
Descriptive characteristics of hospitalizations of patients with a spinal cord infarction.

### Outcomes Overall

Index hospitalizations of patients with a SCI diagnosis had an inpatient morality rate of 14.66% (95% CI: 13.74-15.59). The average LOS for hospitalizations of patients with a SCI diagnosis was 13.09 days (95% CI: 12.61-13.59) and the average hospital cost was $53,531 (95% CI: 51,615-55,518). Most patients were transferred to a skilled nursing facility, intermediate care facility, or another type of facility (48.50%, 95% CI: 47.00-49.99). Relatedly, 16.86% (95% CI: 15.78-17.94) of hospitalizations of patients with a SCI diagnosis resulted in a routine discharge and 3.75% (95% CI: 3.26-4.25) hospitalizations resulted in a transfer to a short-term hospital. For all-cause readmissions, 21.08% (95% CI: 19.86-22.30) and 34.01% (95% CI: 32.36-35.65) of hospitalizations of patients with a SCI diagnosis were readmitted within 30 days and 90 days, respectively.

### Outcomes by Age and Comorbidity Burden

Figure 1 presents the effect of age and CCI on inpatient mortality and LOS. Age was statistically associated with inpatient mortality and LOS (p < 0.001, p < 0.001, respectively); as age increased the rate of inpatient mortality increased (Figure 1A) and LOS decreased (Figure 1B). CCI was statistically associated with inpatient mortality and LOS (p < 0.001, p < 0.001, respectively); as CCI increased the rate of inpatient mortality and LOS increased (Figure 1C-D).

**Figure 1.**
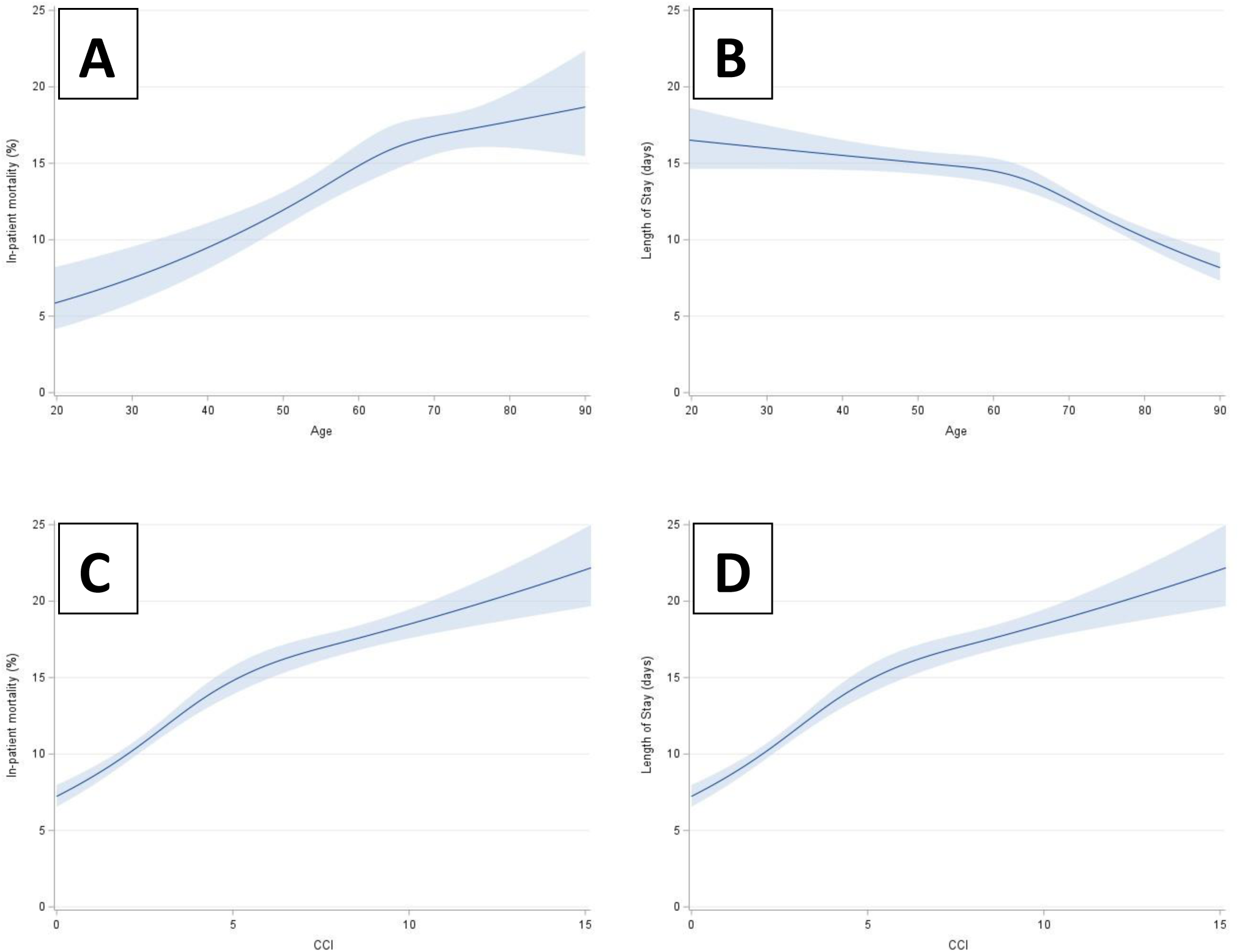
The effect of age (top row) and comorbidity burden (bottom row) on inpatient mortality (left column) and length of stay (right column) for hospitalizations of patients with a spinal cord infarction.

### Outcomes by Thrombolysis

Notably, 1.17% (Weighted N: 147, 95% CI: 111-183) of hospitalizations with a SCI diagnosis received thrombolysis. Hospitalization outcomes by whether patients with SCI received thrombolysis are presented in **Table 2**. Inpatient mortality, length of stay, transfer to a skilled nursing facility, intermediate care facility, or another type of facility, and all-cause 30-day readmission were statistically similar between hospitalization with thrombolysis and hospitalization without thrombolysis. After adjusting for age and CCI, the adjusted odds of 90-day all-cause readmissions were 65% lower for index hospitalizations with thrombolysis compared to no thrombolysis (aOR 95% CI: 0.16-0.77, p = 0.009, **Table 2**).

**Table 2.**
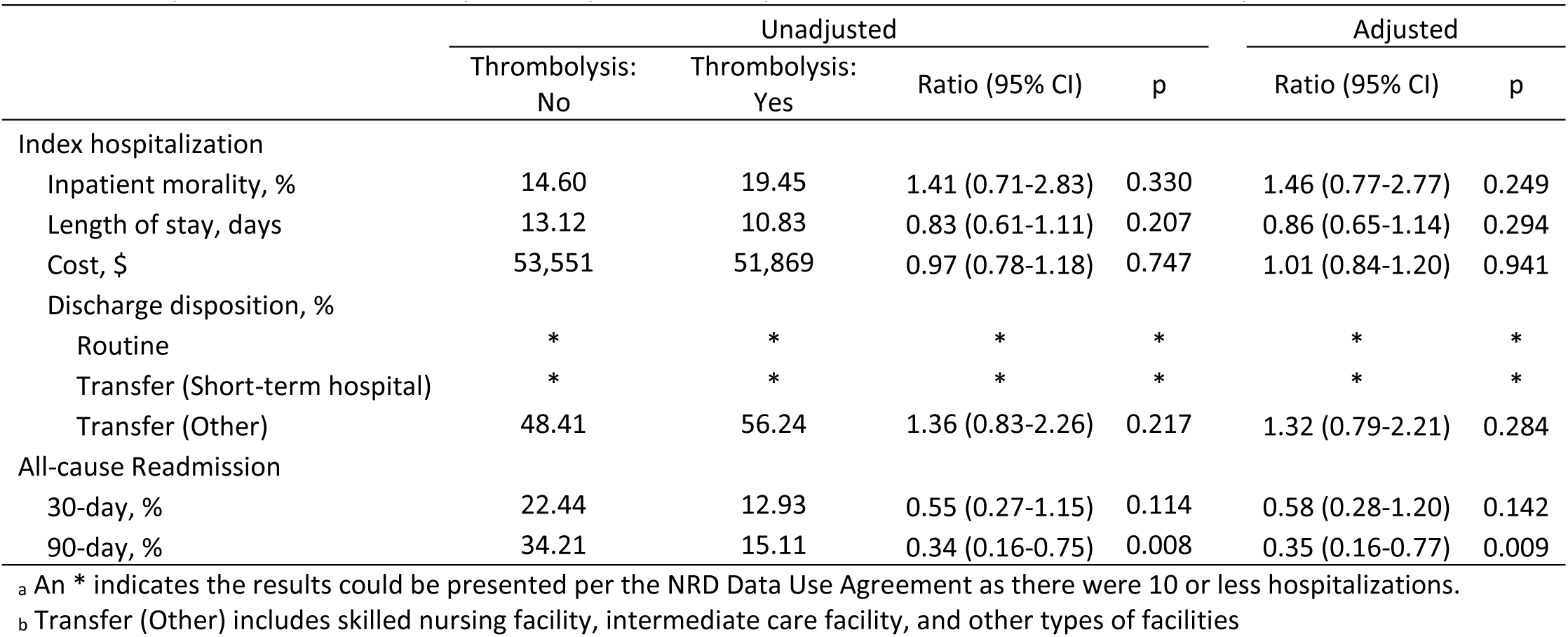
Hospitalizations outcomes by whether patients with spinal cord infarctions received thrombolysis.

### Outcomes by Dissections and Aneurysms

From 2016-2021, 29.12% of hospitalizations of patients carrying a diagnosis for SCI also carried a diagnosis of a dissection or aneurysm (Weighted N: 3,654, 95% CI: 3,393-3,915). **Table 3** presents hospitalization outcomes by whether patients with SCI had an aneurysm or dissection. After adjusting for age and CCI, the adjusted odds of inpatient mortality were 52% greater for SCI hospitalizations of patients with an aneurysm or dissection compared to those without an aneurysm or dissection (aOR 95% CI: 1.30-1.78, p < 0.001, **Table 3**). Likewise, adjusted LOS was 19% greater and adjusted cost was 2.18x greater for SCI hospitalizations of patients with an aneurysm or dissection compared to those without an aneurysm or dissection (LOS-aOR 95% CI: 1.09-2.29, p < 0.001; Cost-aOR 95% CI: 2.06-2.31, p < 0.00; **Table 3**). After adjustment, discharge disposition and readmissions were similar between SCI hospitalizations with and without an aneurysm or dissection (**Table 3**).

**Table 3.** Hospitalization outcomes by whether patients with a spinal cord infarction suffered from an aneurysm or dissection.

|  | Unadjusted |  |  |  | Adjusted |  |
| --- | --- | --- | --- | --- | --- | --- |
|  | No | Yes | Ratio (95% CI) | p | Ratio (95% CI) | p |
| Index hospitalization |  |  |  |  |  |  |
| Inpatient mortality, % | 12.48 | 19.98 | 1.75 (1.51-2.04) | < 0.001 | 1.52 (1.30-1.78) | < 0.001 |
| Length of stay, days | 12.35 | 15.08 | 1.22 (1.12-1.33) | < 0.001 | 1.19 (1.09-1.29) | < 0.001 |
| Cost, \$ | 42,558 | 93,643 | 2.20 (2.07-2.33) | < 0.001 | 2.18 (2.06-2.31) | < 0.001 |
| Discharge disposition, % |  |  |  |  |  |  |
| Routine | 18.57 | 12.69 | 0.64 (0.54-0.75) | < 0.001 | 0.88 (0.74-1.05) | 0.154 |
| Transfer (Short-term hospital) | 3.70 | 3.90 | 1.06 (0.80-1.40) | 0.704 | 1.14 (0.85-1.52) | 0.379 |
| Transfer (Other) | 48.45 | 48.61 | 1.01 (0.90-1.13) | 0.914 | 0.89 (0.79-1.00) | 0.057 |
| All-cause Readmission |  |  |  |  |  |  |
| 30-day, % | 19.95 | 24.13 | 1.28 (1.10-1.48) | 0.002 | 1.16 (0.99-1.36) | 0.067 |
| 90-day, % | 32.88 | 37.04 | 1.20 (1.03-1.40) | 0.017 | 1.06 (0.90-1.24) | 0.472 |

<sup>a</sup> An \* indicates the results could be presented per the NRD Data Use Agreement as there were 10 or less hospitalizations.
<sup>b</sup> Transfer (Other) includes skilled nursing facility, intermediate care facility, and other types of facilities

### Outcomes by Sepsis

Of the estimated 12,548 SCI hospitalizations, 22.23% also had a sepsis diagnosis (Weighted N: 2,789, 95% CI: 2,618-2,959). Hospitalization outcomes by whether patients with SCI also suffered from sepsis are presented in **Table 4**. After adjusting for age and CCI, SCI hospitalizations with sepsis had 3.15x greater adjusted odds of inpatient mortality compared to hospitalizations without sepsis (aOR 95% CI: 2.64-3.75, p < 0.001; **Table 4**). SCI hospitalizations with sepsis had 44% longer adjusted LOS and 54% greater adjusted hospital cost compared to those without sepsis (LOS-aOR 95% CI: 1.34-1.55, p < 0.001; Cost-aOR 95% CI: 1.45-1.65, p < 0.001; **Table 4**). Compared to hospitalization without sepsis, hospitalizations with sepsis had 59% and 33% lower adjusted odds of routine discharge and transfer to a short-term hospital, respectively (Routine-aOR 95% CI: 0.33-0.52, p < 0.001; Short-term Hospital Transfer-aOR 95% CI: 0.48-0.93, p = 0.015; **Table 4**). SCI hospitalizations of patients with sepsis had 42% and 48% greater adjusted odds of 30-day and 90-day all-cause readmission compared to those without a sepsis diagnosis, respectively (30-day-aOR 95% CI: 1.19-1.70, p < 0.001; 90-day-aOR 95% CI: 1.23-1.77, p < 0.001; **Table 4**).

**Table 4.** Hospitalization outcomes by whether patients with a spinal cord infarction suffered from sepsis.

|  | Unadjusted |  |  |  | Adjusted |  |
| --- | --- | --- | --- | --- | --- | --- |
|  | Sepsis:<br>No | Sepsis<br>: Yes | Ratio (95% CI) | p | Ratio (95% CI) | p |
| Index hospitalization |  |  |  |  |  |  |
| Inpatient mortality, % | 22.22 | 25.00 | 1.17 (0.65-2.08) | 0.599 | 3.15 (2.64-3.75) | < 0.001 |
| Length of stay, days | 11.59 | 20.06 | 1.73 (1.61-1.86) | < 0.001 | 1.44 (1.34-1.55) | < 0.001 |
| Cost, \$ | 46,721 | 86,215 | 1.85 (1.73-1.97) | < 0.001 | 1.54 (1.45-1.65) | < 0.001 |
| Discharge disposition, % |  |  |  |  |  |  |
| Routine | 19.33 | 8.22 | 0.37 (0.30-0.47) | < 0.001 | 0.41 (0.33-0.52) | < 0.001 |
| Transfer (Short-term hospital) | 3.99 | 2.92 | 0.72 (0.53-0.99) | 0.044 | 0.67 (0.48-0.93) | 0.015 |
| Transfer (Other) | 48.50 | 48.51 | 1.00 (0.88-1.14) | 0.993 | 1.00 (0.87-1.14) | 0.975 |
| All-cause Readmission |  |  |  |  |  |  |
| 30-day, % | 19.42 | 28.43 | 1.65 (1.39-1.96) | < 0.001 | 1.42 (1.19-1.70) | < 0.001 |
| 90-day, % | 31.74 | 44.21 | 1.70 (1.43-2.02) | < 0.001 | 1.48 (1.23-1.77) | < 0.001 |

An \* indicates the results could be presented per the NRD Data Use Agreement as there were 10 or less hospitalizations.
Transfer (Other) includes skilled nursing facility, intermediate care facility, and other types of facilities

## Discussion

Given the rarity of SCI, previous studies have been limited by the number of patients^2,3^. Our study analyzed 12,548 SCI hospitalizations from 2016 to 2021 using the NRD. Most SCI hospitalizations were in large and metropolitan teaching hospitals. Diagnosing SCI is not an easy feat, and most cases require higher levels of care and specialized care^2^. In our study, there were an estimated 147 SCI hospitalizations where the patient received thrombolysis. Notably, hospitalizations where the patient received thrombolysis had significantly lower odds of 90-day all-cause readmissions compared to hospitalizations where the patient did not receive thrombolysis. This difference remained even after adjusting for age and CCI. However, there was no significant difference in the rate of inpatient mortality or 30-day all-cause readmissions between hospitalizations with thrombolysis and hospitalizations without thrombolysis. Studies have previously shown that recovery after spinal cord infarction can take several months^2,11^. It is, therefore, plausible that thrombolysis might help in the long-term recovery after SCI. A study using a national registry in Denmark showed that thrombolysis in cerebral stroke had lower readmission rates for pneumonia, but no overall effect on readmission rates^21^. On the other hand, Vahidy et al. analyzed the NRD for cerebral strokes and found significantly lower odds of 30-day readmission with recanalization therapy which included thrombolysis and thrombectomy^29^. Also, previous work on cerebral stroke showed the long-term benefits of thrombolysis in terms of functional outcome and independence several months after treatment^22,23^. However, thrombolysis in cerebral stroke has been shown to be associated with higher in-hospital mortality^24^. This is also compatible with our findings of 19.45% inpatient mortality of hospitalizations with thrombolysis compared to 14.60% for hospitalizations without thrombolysis. However, the difference in mortality was not statistically significant. Our study shows an association between thrombolysis and better long-term outcomes in SCI. However, large trials are needed to establish the benefit and risk of thrombolysis in SCI with detailed functional outcomes.

Hospitalizations specifically for SCI were the primary reason patients with a SCI diagnosis were hospitalized; interestingly the other most common reasons a patient with a SCI diagnosis was hospitalized were aneurysms and dissections followed by sepsis. Hospitalizations primarily for an aneurysm or dissection had significantly higher inpatient mortality, longer hospital stays, and higher overall costs compared to hospitalizations where the patient with SCI did not experience an aneurysm or dissection. These are expected findings given that aortic pathology is one of the most common conditions leading to SCI^25^. Aortic aneurysms and dissections are also likely to require surgical intervention, leading to worse outcomes overall^26,27^. These findings carry prognostic value in patients who suffer from SCI, regardless of age or comorbidity burden. Among all the hospitalizations with SCI, there were no SCI hospitalizations where the patient had a percutaneous aortic repair. There were 68 SCI hospitalizations where the patient had an open surgical aortic repair. This finding is consistent, yet more remarkable, when compared to previous studies showing lower rates of peri-procedural SCI after percutaneous aortic repairs compared to open aortic repairs^8^.

We found similar outcomes in hospitalizations that were primarily for sepsis; sepsis was associated with longer hospital stays and greater costs. The odds of inpatient mortality were greater for SCI hospitalizations with concomitant sepsis compared to those without sepsis. Sepsis disproportionately affects older patients with medical comorbidities^30^. Previous studies have shown that sepsis accounted for 28% of inpatient mortality in spinal cord injuries^28^. The association between sepsis and SCI is bidirectional. Septic shock can potentially lead to hypoperfusion and spinal cord ischemia, and septic emboli are known to cause cerebral stroke^31^. Alternatively, it is plausible that SCI is commonly complicated by sepsis. These findings may highlight the important role sepsis plays in determining inpatient mortality in SCI. Furthermore, all-cause readmission rates at 30- and 90-days were significantly higher in SCI hospitalizations with sepsis. Studies that examined admissions solely for sepsis found high rates of readmissions with most of them occurring within 90 days^32^.

We found that as age increased, inpatient mortality increased, contributing to shorter length of stays and lower costs found in the older age groups of SCI hospitalizations. The median age in our study was 63 (53-73), which is slightly higher compared to previous reports^3^. Comorbidity burden played an important role in inpatient mortality and LOS for SCI hospitalization; comorbidity burden was significantly associated with inpatient mortality and LOS. This is consistent with reports on cerebral infarction. Han et al. reported a significantly poorer prognosis in ischemic strokes with a CCI of 3 or more^33^. Our study showed a median CCI of 2 with 50% of hospitalizations being patients with a CCI between 3 and 7 thus indicating the high comorbidity burden in patients with SCI. In accordance with published reports, we found that peripheral vascular disease was the most common comorbidity in our SCI hospitalizations^3,4^. Interestingly, AIDS was one of the common comorbidities in SCI hospitalizations (21.46%). Anecdotal evidence exists to suggest an association between opportunistic infections, such as Varicella Zoster Virus, and vasculitis leading to SCI^34^. This finding warrants further investigation into the association between AIDS and SCI.

The NRD is an administrative database reliant on billing codes, non-reimbursable codes may be underreported. However, the NRD is still a useful database to examine uncommon conditions, as it captures a large number of hospitalizations on a national and representative scale. The NRD does not provide information on long-term functional outcomes. Our analysis on thrombolysis is limited by the low number of hospitalizations, due to possible under-reporting of thrombolysis.

## Conclusions

This study highlights the factors that impact the outcomes of SCI on a large scale. We found that age, comorbidity burden, and concomitant aneurysms, dissections and sepsis all play major roles in the outcome of SCI. These findings can assist in evaluating prognosis after SCI. Additionally, the findings uncover the association between thrombolysis and better long-term outcomes in SCI. This paves the way for further investigation into the role of thrombolysis in SCI. Large randomized controlled trials are needed to establish the benefit and risk of thrombolysis in SCI.

## Data Availability

The Nationwide Readmissions Database is HIPAA-compliant and publicly available. Data directly used for this study can be made available upon request to the corresponding author.

## Acknowledgements and Sources of Funding

None.

## Conflicts of Interest, Consent and IRB

We have no conflicts of interest to disclose. Creighton University acknowledged this study as Not Human Subjects Research (InfoEd Record Number: 2004608); the NRD is HIPAA-compliant and publicly available. Therefore, informed consent was deemed unnecessary for this retrospective analysis of de-identified data.

## Author Contributions

Ali Al-Salahat: Conception/Design, methodology, literature review, writing manuscript/critical revision

Danielle Dilsaver: Design, Statistical Analysis, Writing Manuscript/Critical revision Ali Bin Abdul Jabbar, Conception/Design, Critical revision

Abhishek Singh: Conception/ Design, Critical revision

